# Benefit take-up in the last year of life: a population-based study using linked data for England and Wales

**DOI:** 10.64898/2026.04.10.26350614

**Authors:** Joanna M Davies, Amie Fairs, Daniel Ayoubkhani, Juliet Stone, Steve Marshall, Melanie Diggle, Andy Bradshaw, Maddy French, Jamilla Hussain, Geoff Fimister, Richard Harding, Katherine E Sleeman, Vahé Nafiliyan

## Abstract

**Context:** In the UK, and in other countries, people living with a terminal illness are eligible for financial support to help with the costs of serious illness and to support their dignity and independence. This study investigates the take-up of benefits in the last year of life and identifies sociodemographic, clinical, and geographical factors associated with underclaiming.

**Methods:** Retrospective cohort study using linked mortality, Census and benefits data for all people who died aged 16+ from chronic illnesses in England and Wales between 1 May 2018 and 30 April 2021. Outcome was receipt of non-means tested disability benefits in the last 12 months of life. We describe geographical variation in take up, and association with sociodemographic, clinical and geographical exposures using Poisson models.

**Findings:** Our population included 1,049,493 eligible decedents, with an overall take-up rate of 65.9%. After adjusting for sociodemographic factors, variation in take-up by cause of death was wide: liver disease 44% (95% CI 43–45%), heart failure 52% (51–52%), cancer 62% (61–62%), dementia 75% (74–75%), and neurodegenerative diseases 90% (88–91%). Across Local Authorities, the age-and-sex-standardised take-up varied from 53% to 78%; rates were generally higher in more deprived areas, but not uniformly.

**Conclusions:** In England and Wales, 1 in 3 people who die from expected causes (120,000 each year) do not receive the benefits for which they are eligible. Our analysis uses novel data linkages and highlights clinical and sociodemographic groups and geographical areas that could be targeted with proactive take-up initiatives.

## Background

Worldwide, 62 million people die each year.^1^ Populations globally are ageing and there is an increase in the number of deaths from non-communicable diseases such as cancer, dementia, cerebrovascular disease, and lung disease.^2^ Chronic diseases are projected to account for 47% of all deaths worldwide by 2060.^2^ Having a terminal illness puts people under significant financial strain through additional costs and loss of earnings for patients and their carers.^3–5^ People living with terminal illness often face increased household bills as they spend more time at home and may have additional costs such as heating, social care, equipment, and transport.^6^ In the United Kingdom (UK) an estimated 1 in 6 people die in poverty (111,000 each year).^7^ Improving access to social security benefits for people with a terminal illness has the potential to alleviate some of the effects of financial hardship and improve dignity and choice towards the end of life.

People living with a terminal illness are eligible for fast-tracked access to social security benefits in the UK^8^ and in other countries including the United States^9^, Canada^10^, and Australia^11^. In England, Wales and Northern Ireland, the ‘Special Rules for End of Life’ apply to people who are expected to be in the last year of life (extended from a 6-month prognosis, a rule enacted between 2022 and 2023).^12^ To access benefits through the ‘Special Rules’, a doctor or specialist nurse provides medical evidence of their patient’s illness, and this expedites access to specific disability, unemployment, and low-income benefits without the need for further medical assessment.^8^ This includes the non-means tested disability benefit Personal Independence Payment (PIP) (currently (2026-7) worth up to

£194.60 per week depending on mobility needs) for working-age people, and Attendance Allowance (AA) (currently worth up to £114.60 per week) for people of state pension age and above. Everyone aged 16 years and over who is recognised by a doctor or specialist nurse as being likely to be in the last year of their life is eligible for either PIP or AA, regardless of their income or assets. People with a terminal illness living on a low income are also eligible for fast-track access to means-tested benefits such as Universal Credit or Pension Credit, designed to help with living costs.

The purpose of fast-tracking access to benefits for people in the last year of life is to ensure rapid access to essential financial support to alleviate some of the costs of serious illness and to help people maintain their independence and quality of life.^6^ Non-take-up of benefits (i.e. when people do not take-up a benefit that they are eligible for), undermines the effectiveness of social protection programmes. It may also increase health and social care costs in the long term if for example people are less able to maintain their own care at home as a result of not receiving benefits.^13^ There are many reasons why people may not take up the benefits they are entitled to, including perceived ineligibility, barriers or lack of support in the system, stigma, or concerns about how claiming could affect other benefits.^6, 14, 15^ The take-up rate varies by benefit type, geographical location and over time, and there is no single methodology for measuring benefit take-up.^16^ A key challenge is that the number of eligible people is not always known to the agencies making the payments, thus estimates often draw on survey data.^16, 17^ A recent study used a combination of administrative and survey data to estimate disability benefit eligibility and take-up for blind and partially sighted people in the UK. It found that 1 in 4 were not claiming the benefits to which they were entitled, and that people in low-income households, of working-age, and from ethnic minority groups were significantly less likely to claim.^18^

To our knowledge, no-one has studied how many people take up the benefits available to them in the last year of life, or how this might vary across England and Wales. To improve benefit take-up for people living with terminal illness, we need a better understanding of variation in the take-up rate and to identify groups most likely to under-claim. Therefore, the aim of this study is to determine the take-up of non-means tested benefits for people in the last year of life and to identify sociodemographic, clinical, and geographical factors associated with underclaiming.

## Methods

### Study design, data sources, setting and participants

This is an observational retrospective cohort study using i) death registrations linked to 2011 and 2021 Census data, from the Office for National Statistics (ONS), and also linked to ii) benefits claims data (Benefit and Income Data Set (BIDS)) from the UK Government Department for Work and Pensions (DWP). Census responses were linked to death registrations using the NHS Patient Register 2011-2013 and the NHS Personal Demographics Service 2019,^19, 20^ and were linked to the benefit data via the Demographic Index,^21, 22^ enabling linkage across all datasets. Data were deidentified prior to analysis, and all analysis was carried out in a secure environment at the ONS.

The study population includes all deaths that occurred between 1^st^ May 2018 and 30^th^ April 2021 that were registered by 31^st^ December 2023 among people aged 16 years and older in England and Wales, who were enumerated in the 2011 or 2021 Census (linkage to the 2011 Census was prioritised, the 2021 Census was only used for deaths occurring on or after 21^st^ March 2021 [i.e. the day of Census 2021] if no Census 2011 record could be linked), and had an encrypted National Insurance number that could be linked to the benefit data. The study time period reflects the most recent benefit and income data available at the time of analysis. The minimum age of 16 years old reflects the minimum age that people become eligible for working age benefits.

We investigated take-up among people who died from chronic progressive illness where there is a reasonable expectation that a healthcare professional could anticipate they were approaching the end of life and thus be eligible to access benefits under the ‘Special Rules for End of Life’. Therefore, we restricted the analysis to people who died from one of seven leading chronic illnesses (following an established classification method,^23, 24^ detailed in table 1), excluding people who died from other causes such as external, sudden and other non-chronic illnesses.

**Table 1:** Exposure variables.

| **Variable name and source** | **Details** |
| --- | --- |
| **Age at death**<br>Source: ONS death registrations | In years at last birthday and as a binary indicator:<br>• working age (16 to 65 years)<br>• pension age ($\geq 66$ years) |
| **Sex**<br>Source: Census 2011 or 2021 | Binary categories:<br>• male<br>• female |
| **Ethnicity**<br>Source: Census 2011 or 2021 | In ten categories:<br>• White British<br>• White other (including Irish, Traveller, any other White background)<br>• Mixed/Multiple ethnic groups (including White and Black Caribbean, White and Asian, White and Black African, and any other mixed background)<br>• Bangladeshi<br>• Indian<br>• Pakistani<br>• Chinese<br>• Black African<br>• Black Caribbean<br>• Other ethnic group (including Arab, any other Asian, any other Black and any other ethnic background) |
| **Relative level of area-based deprivation**<br>Source: IMD (for England), WIMD (for Wales), linked to the LSOA code of the participant's usual residence recorded at the Census 2011 or 2021 | In national quintile groups (1 is most deprived), separately for England and Wales.<br>Area-based deprivation was used as a proxy for individual-level socioeconomic position and based on the Index of Multiple Deprivation for England (IMD) 2019, and the Welsh Index of Multiple Deprivation (WIMD) 2019, linked at the Lower Super Output Area (LSOA) (neighbourhood) level to the participant's postcode at the Census.<sup>29, 30</sup> |
| **Highest level of education**<br>Source: Census 2011 or 2021 | In seven categories (see supplementary file, p1 for further details of the qualifications included in each category):<br>• no qualifications<br>• level 1 (1-4 GCSEs etc)<br>• level 2 (5+ GCSEs etc)<br>• apprenticeships<br>• level 3 (2+ A levels etc)<br>• level 4 (degree etc)<br>• other qualifications (vocational and foreign) |
| **English or Welsh language proficiency**<br>Source: Census 2011 or 2021 | Binary categories:<br>not well/not at all<br>very well/well/native speaker |
| **Region**<br>Source: linked to the LSOA code of the participant's usual residence recorded at the Census 2011 or 2021 | Participant's region at the Census, including nine regions of England, and Wales |
| **Local Authority**<br>Source: linked to the LSOA code of the participant's usual residence recorded at the Census 2011 or 2021 | Participant's Lower Tier Local Authority (LA) at the Census mapped on to 2021 LA geography, including 307 LAs in England (due to small counts, we grouped City of London with Hackney, and Isles of Scilly with Cornwall), and 22 in Wales |
| **Underlying cause of death**<br>Source: ONS death registrations | In seven categories of chronic illness, defined following an established method for identifying expected causes of death,<sup>23, 24</sup> using the International Statistical Classification of Diseases and Related Health Problems 10th Revision (ICD-10) codes.<br>Cancer (C00-C97)<br>Dementia/Alzheimer's/Frailty (F01, F03, G30, G31, R54)<br>Heart disease and heart failure (I00-I52 (excl. I12 & I13))<br>Chronic lower respiratory disease, respiratory failure (J40-J47, J96)<br>Reno-vascular disease, renal failure (I12, I13, N17, N18, N28)<br>Liver disease (K70-K77)<br>Other chronic illness including:<br>Neurodegenerative (Huntington's, MND, Parkinson's, PSP, MS, MSA) (G10, G12.2, G20, G23.1, G35, G90.3)<br>Stroke (I60-I69)<br>HIV (B20-B24)<br>Excluding all other underlying causes of death not listed above, such as external, sudden and other non-chronic illnesses. |

ONS: Office for National Statistics

### Variables

The outcome variable is a binary indicator of whether the decedent received one of three non–means-tested disability benefits at any point during the last 12 months of life, including Personal Independence Payment (PIP) and Disability Living Allowance (DLA) for working-age people, and Attendance Allowance (AA) for people of pension age. PIP gradually replaced DLA from 2013 onwards; however, some individuals have continued to receive DLA.^25^

The Department for Work and Pensions (DWP) record the main disabling condition associated with each disability benefit claim, including whether the claim was made under the ‘Special Rules’ for a terminal illness. However, this information was not available in the data used in this study. Therefore, our outcome measure does not indicate whether individuals claimed disability benefits under the ‘Special Rules’, but rather whether they received a non–means-tested disability benefit for any disabling condition during their last year of life. Non-receipt among those who died from an expected cause indicates missed access to non–means-tested disability benefits for which they would have been eligible under the ‘Special Rules’.

People who enter state-funded residential or nursing care, or who have a hospital admission lasting 28 days or more, may have their disability benefit payments suspended or stopped in line with payment rules. To avoid underestimating benefit take-up—particularly among older adults, who are more likely to enter state-funded institutional care—we disregarded benefit ‘exits’ prior to death and instead defined benefit receipt as receipt at any point during the final 12 months of life.

The study period predates the 2023 policy change that extended eligibility for PIP and AA under the ‘Special Rules’ from a six-month to a 12-month prognosis. We chose to focus on the last year of life to align with current policy and with research and clinical practice, which commonly focus on the last year of life. The 12-month period also helps to minimise underestimation of benefit receipt due to benefit ‘exits’ prior to death.

Exposure variables (detailed in table 1) were selected based on existing evidence on the social, clinical, and geographical factors known to be important to accessing care and support towards the end of life,^26–28^ and on availability of variables in the datasets.

### Analysis

#### Cumulative take-up over the last 12 months of life

We describe and graphically depict the cumulative proportion of people claiming non-means tested benefits over the last 12 months of life, separately for people of working and pension age.

#### Descriptive analysis

We describe the number and proportion of people claiming non-means tested benefits in the last year of life overall, and by the different exposure variables.

#### Geographical analysis

Using the direct method of standardisation^31^, we present the age and sex standardised take-up rate for non-means tested benefits in the last year of life for each Lower Tier Local Authority (LA) in England and Wales and visually depict this in a map. The total number of deaths from chronic illnesses in the study population overall for England and Wales combined was used as the standard population, stratified into the following age bands, separately for men and women: ≤20 years, 10-year age bands from 21-30 to 91-100, and >100.

We plot the association between the level of take-up and the relative level of deprivation in each LA using a scatter plot and use linear regression to model the association between the two variables. The LA deprivation score was calculated as the population-weighted average of the combined IMD scores for the Lower Super Output Areas (LSOAs, i.e. neighbourhoods) in each LA; higher scores represent more deprived areas (NB this contrasts with the direction of the coding for the IMD deprivation quintile groups where quintile 1 is most deprived and quintile 5 is least deprived).^29^

#### Modelling

We modelled the binary outcome (take-up or not of non-means tested benefits in the last year of life) using Poisson models with robust standard errors to account for heteroscedasticity. We chose Poisson models because risk ratios are arguably more easily interpreted than odds ratios.^32^ We did not have a main exposure of interest, rather we had a set of candidate exposures and thus, we present a model for each exposure minimally adjusted for age and sex, and a model fully adjusted for all exposure variables. Models were restricted to cases with complete data across all variables. Age was treated as a numerical variable and modelled using a natural spline to account for non-linearity in the relationship between age and the outcome. Spline parameters were determined after testing combinations of internal and boundary knot settings and choosing the model with the lowest Bayesian Information Criterion (BIC) value.

To help interpret the adjusted associations, we report predicted proportions of the benefit take-up rate, calculated using the emmeans package in R.^33^ The predicted proportions were estimated using proportional weights and are therefore interpreted at the mean age of the study population and at the average value of the categorical covariates in the model.

To understand how the associations might vary according to age, we interacted the exposures with age as a binary variable, splitting the study population into working age (16-65 years) and pension age (≥66 years). The interaction models were adjusted for age (modelled as a natural spline) and sex, and the predicted proportions were calculated with proportional weights.

Point estimates are accompanied by 95% confidence intervals (CIs), and statistical significance was determined at the 5% level (p<0.05). Data preparation was conducted using Sparklyr version 1.92^34^ and all statistical analyses were performed using R version 4.4.^35^

## Results

Our study population included 1,049,493 people aged over 16 years old who died of a chronic illness in England and Wales between 1st May 2018 and 30th April 2021. We started with all deaths of people resident in England and Wales over the study period (1,690,146) and excluded 193,496 (12%) deaths during the linkage process (figure 1 provides the reasons for these exclusions). Descriptive data on the 446,067 people who died from causes not included in the main study population such as external injury, sudden and non-chronic illnesses are provided in supplementary file (table 1, 2 and figure 1).

**Figure 1:**
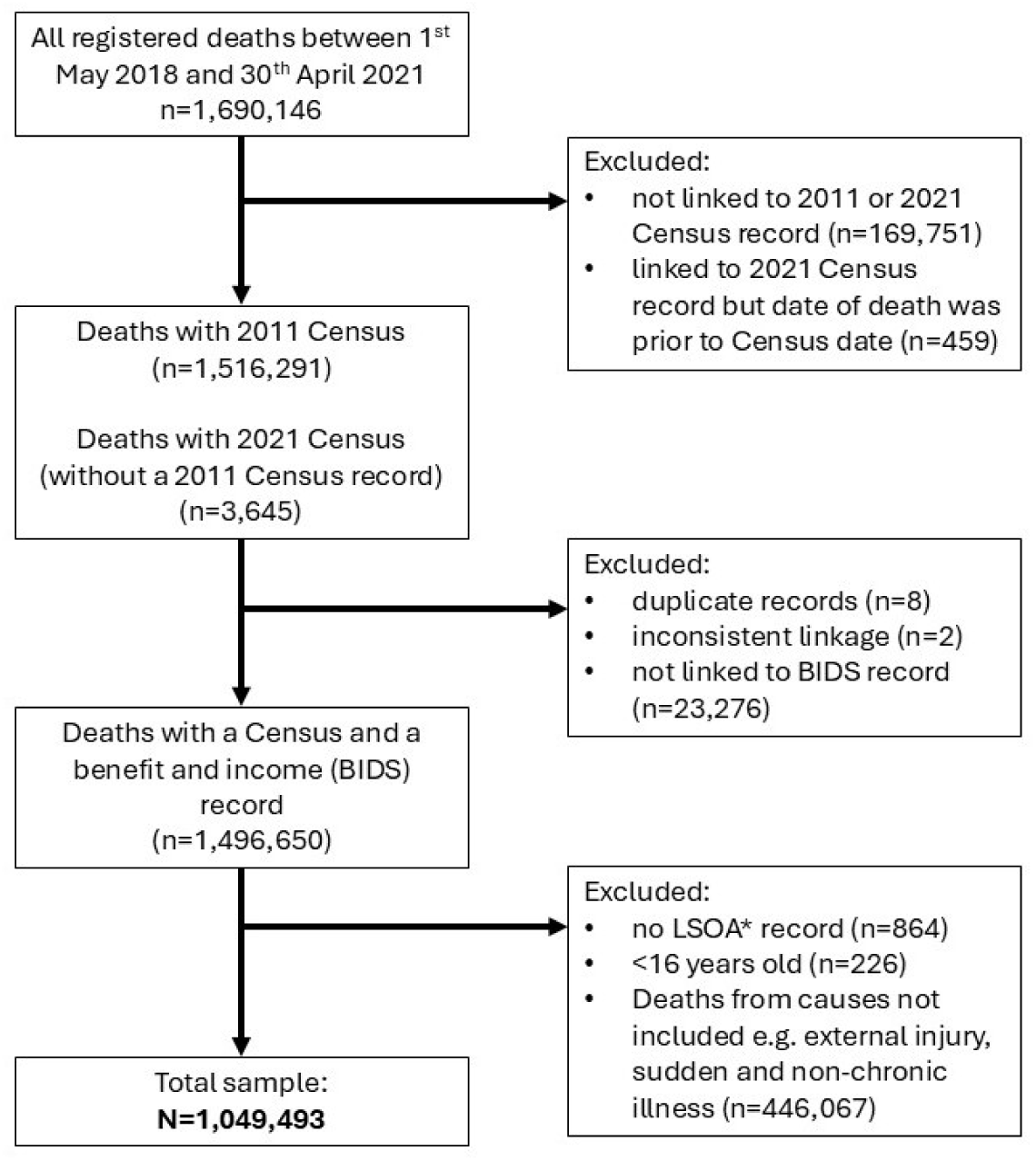
Diagram of study population inclusion and exclusion. *LSOA: Lower Super Output Area (neighbourhood)

The study population of people who died from chronic illness, and variation in the level of benefit take-up across different characteristics are described in Table 2. Overall, 65.9% of decedents received non-means tested benefits in their last year of life.

**Table 2:** Study population descriptives for all people (aged 16+) who died from chronic illnesses in England and Wales between 1st May 2018 and 30th April 2021, and the take-up of non-means tested disability benefits: Personal Independence Payment (PIP), Attendance Allowance (AA) and Disability Living Allowance (DLA)

|  | all deaths<br>from chronic<br>illnesses:<br>count | column % | n receiving<br>non-means-tested<br>benefits in<br>last 12<br>months of<br>life | row %<br>receiving<br>non-means-tested<br>benefits in<br>last 12<br>months of<br>life) |
| --- | --- | --- | --- | --- |
| **Total N** | 1049493 | 100.0% | 691669 | 65.9% |
| **Median age (Interquartile Range)** | 81 (72, 88) |  | 83 (74, 89) |  |
| **Working age at death (16-65)** | 142432 | 13.6% | 78308 | 55.0% |
| Median age (Interquartile Range) | 58 (52, 62) |  | 59 (53, 63) |  |
| **Pension age at death (≥66)** | 907061 | 86.4% | 613361 | 67.6% |
| Median age (Interquartile Range) | 83 (76, 89) |  | 85 (78, 90) |  |
| **Sex** |  |  |  |  |
| Female | 528516 | 50.4% | 375089 | 71.0% |
| Male | 520977 | 49.6% | 316580 | 60.8% |
| **Ethnicity** |  |  |  |  |
| White British | 973750 | 92.8% | 643596 | 66.1% |
| White other | 33337 | 3.2% | 21380 | 64.1% |
| Mixed/Multiple ethnicity | 4559 | 0.4% | 2959 | 64.9% |
| Bangladeshi | 1919 | 0.2% | 1349 | 70.3% |
| Indian | 10822 | 1.0% | 6628 | 61.2% |
| Pakistani | 5216 | 0.5% | 3519 | 67.5% |
| Chinese | 1575 | 0.2% | 809 | 51.4% |
| Black African | 2869 | 0.3% | 1504 | 52.4% |
| Black Caribbean | 8740 | 0.8% | 5866 | 67.1% |
| Other ethnic group | 6436 | 0.6% | 3901 | 60.6% |
| Missing or not stated | 270 | 0.0% | 158 | 58.5% |
| **Area-based deprivation** |  |  |  |  |
| 1 (most deprived) | 203890 | 19.4% | 144680 | 71.0% |
| 2 | 202704 | 19.3% | 136199 | 67.2% |
| 3 | 214872 | 20.5% | 140511 | 65.4% |
| 4 | 217052 | 20.7% | 139124 | 64.1% |
| 5 (least deprived) | 210224 | 20.0% | 130649 | 62.1% |
| Missing or not stated | 751 | 0.1% | 506 | 67.4% |
| **Highest educational qualification** |  |  |  |  |
| No qualifications | 531903 | 50.7% | 381127 | 71.7% |
| Level 1 (1-4 CSEs etc) | 76551 | 7.3% | 45359 | 59.3% |
| Level 2 (5+ CSEs etc) | 90344 | 8.6% | 54795 | 60.7% |
| Apprenticeships | 59888 | 5.7% | 37096 | 61.9% |
| Level 3 (2+ A levels etc) | 52099 | 5.0% | 29956 | 57.5% |
| Level 4 (degree etc) | 176373 | 16.8% | 102251 | 58.0% |
| Other qualifications: Vocational/Work-related/Foreign | 61234 | 5.8% | 40421 | 66.0% |
| Missing or not stated | 1101 | 0.1% | 664 | 60.3% |
| **English/Welsh language proficiency** |  |  |  |  |
| Not well/Not at all | 10056 | 1.0% | 7241 | 72.0% |
| Very well/Well | 15994 | 1.5% | 9690 | 60.6% |
| Native speaker | 1023173 | 97.5% | 674580 | 65.9% |
| Missing | 270 | 0.0% | 158 | 58.5% |
| **Region** |  |  |  |  |
| North East | 54322 | 5.2% | 38090 | 70.1% |
| North West | 143011 | 13.6% | 99223 | 69.4% |
| Yorkshire and The Humber | 105882 | 10.1% | 72111 | 68.1% |
| East Midlands | 91325 | 8.7% | 60460 | 66.2% |
| West Midlands | 109224 | 10.4% | 72988 | 66.8% |
| East of England | 113338 | 10.8% | 72101 | 63.6% |
| London | 97069 | 9.2% | 59563 | 61.4% |
| South East | 159492 | 15.2% | 98917 | 62.0% |
| South West | 109529 | 10.4% | 70580 | 64.4% |
| Wales | 65550 | 6.2% | 47130 | 71.9% |
| Missing | 751 | 0.1% | 506 | 67.4% |
| **Underlying cause of death** |  |  |  |  |
| Cancer | 395307 | 37.7% | 245034 | 62.0% |
| Dementia/Alzheimer's/senility | 201782 | 19.2% | 171937 | 85.2% |
| Heart failure | 239402 | 22.8% | 130089 | 54.3% |
| HIV | 227 | 0.0% | 93 | 41.0% |
| Liver disease | 21686 | 2.1% | 9178 | 42.3% |
| Neurodegenerative diseases | 30063 | 2.9% | 26979 | 89.7% |
| Renal failure | 6657 | 0.6% | 4783 | 71.8% |
| Respiratory failure | 75830 | 7.2% | 55409 | 73.1% |
| Stroke | 78539 | 7.5% | 48167 | 61.3% |

### Cumulative take-up over the last 12 months of life

Figure 2 (and supplementary table 2) shows the cumulative non-means tested benefit take-up rate per 1,000 deaths over the last 12 months of life, separately for working age and pension age people. The take-up rate increased gradually over the last year of life, with a steeper increase in the last 6 months of life particularly for those of working age.

**Figure 2:**
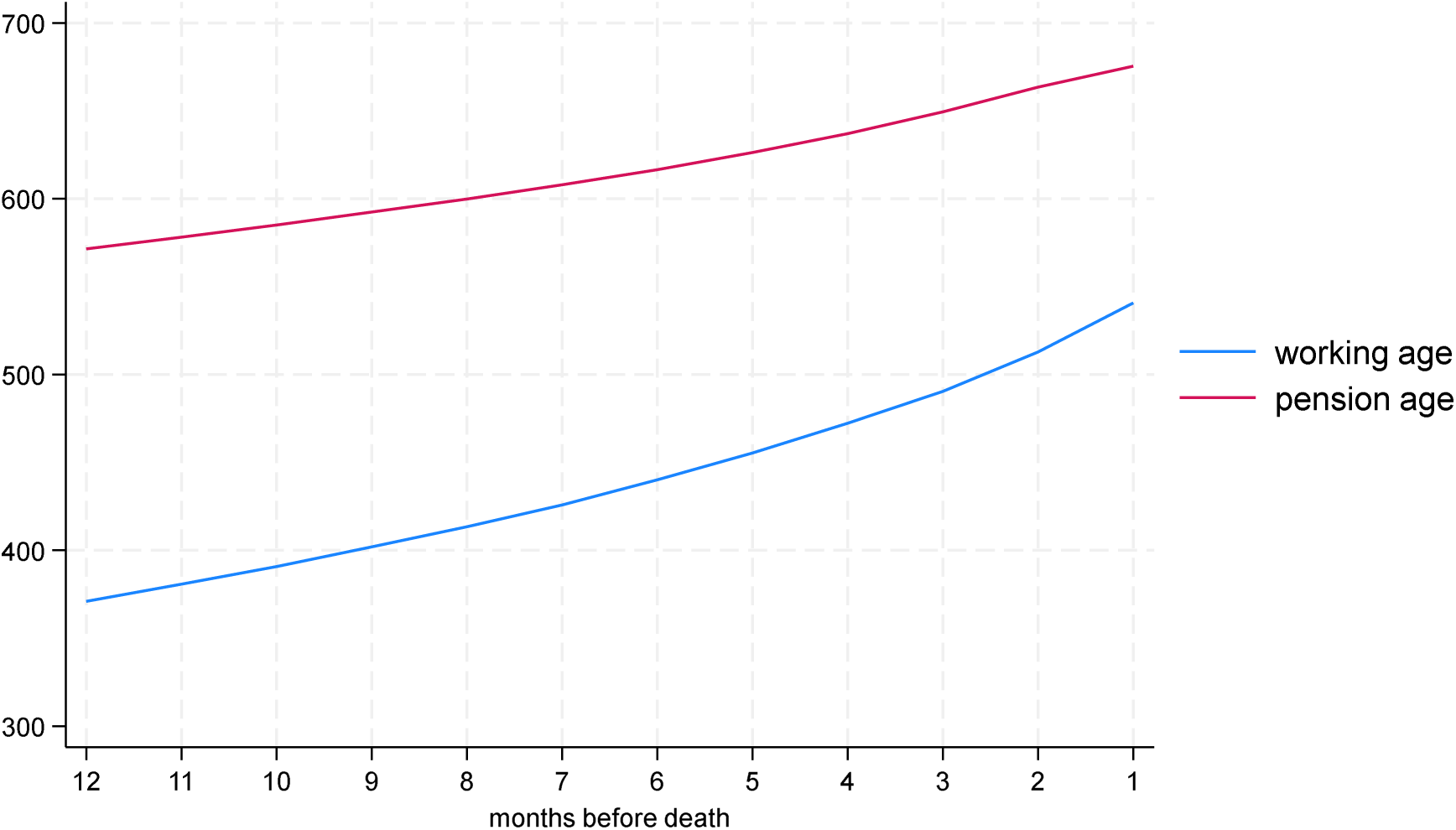
Cumulative take-up rate for non-means tested disability benefits (Personal Independence Payment (PIP), Attendance Allowance (AA) and Disability Living Allowance (DLA)), over the last 12 months of life, per 1000 deaths, for all deaths (aged 16+) from chronic illnesses in England and Wales between 1st May 2018 and 30th April 2021 (n=1,049,493)

#### Geographical analysis

Figure 3 shows the geographical variation in the age and sex standardised benefit take-up rate across England and Wales. The take-up rate across LAs ranged from 78.3% to 52.5% and tended to be higher in Wales and the North of England than in the South of England.

**Figure 3:**
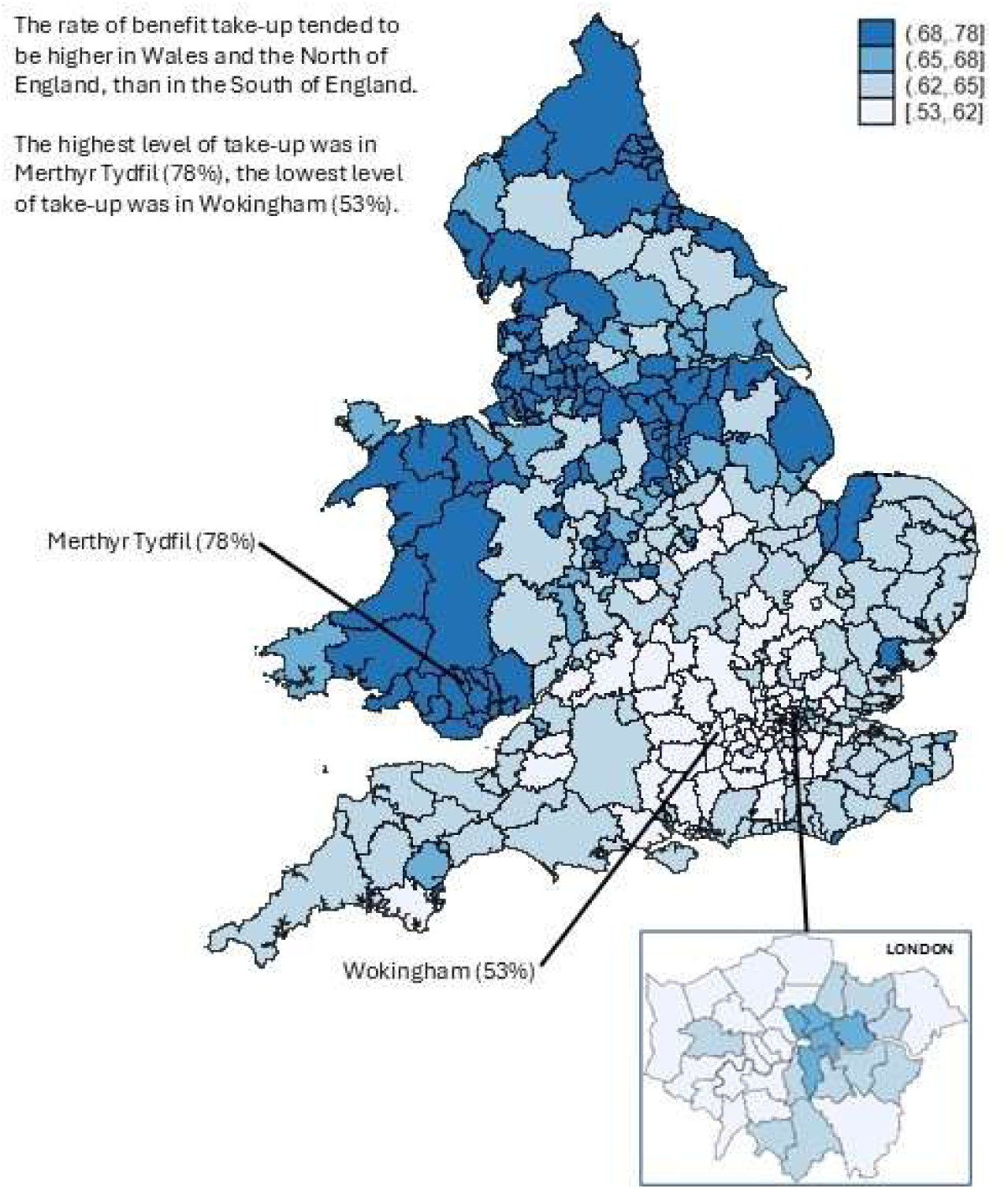
Map of variation in the age and sex standardised benefit take-up rate for non-means tested disability benefits (Personal Independence Payment (PIP), Attendance Allowance (AA) and Disability Living Allowance (DLA)) in Local Authorities in England and Wales (n=329), for all people who died (aged 16+) from chronic illnesses between 1st May 2018 and 30th April 2021 (n=1,049,493)

In England and Wales, there was a positive association between the average deprivation score in each LA (higher scores are more deprived) and the age and sex standardised rate of benefit take-up (Figure 4). Areas with lower benefit take-up rates tended to be less deprived, however, this was not uniform. Among the 20% of LAs with the lowest take-up rate, there were 7 (all in London or the South of England) with higher levels of deprivation, including: Haringey (59% take-up; in 20% most deprived LAs); Kensington & Chelsea (56% take-up; in 40% most deprived LAs); Hammersmith and Fulham (58% take-up; in 40% most deprived LAs); Hounslow (58% take-up; in 40% most deprived LAs); Brent (60% take-up; in 40% most deprived LAs); Slough (61% take-up; in 40% most deprived LAs) and Enfield (61% take-up; in 40% most deprived LAs). See supplementary file table 3 for full details.

**Figure 4:**
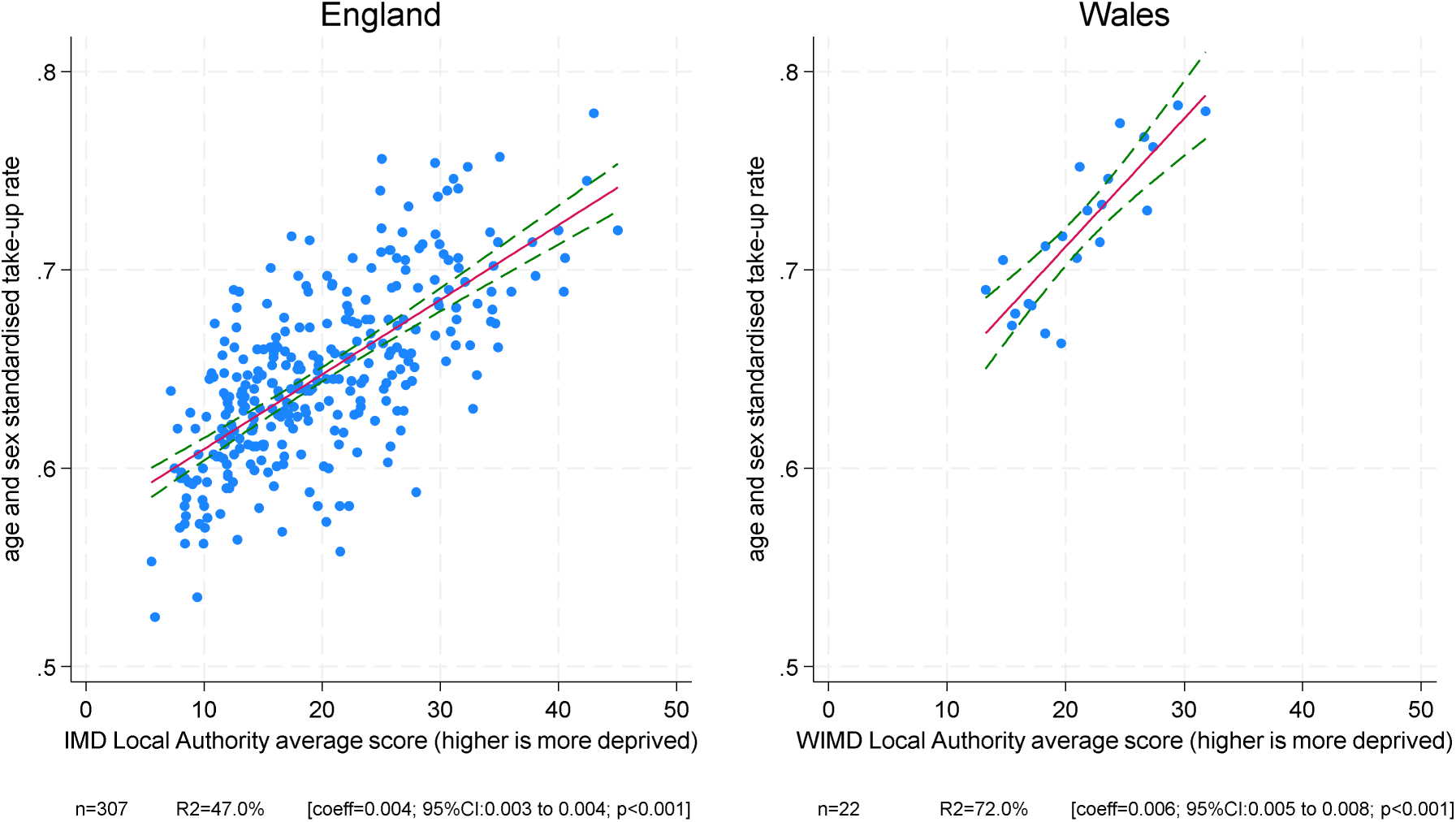
Scatter plot (with linear line of fit and 95% CI) of the age and sex standardised take-up rate for non-means tested disability benefits (Personal Independence Payment (PIP), Attendance Allowance (AA) and Disability Living Allowance (DLA)), against average deprivation in each Local Authority in England and Wales, for all people who died (aged 16+) from chronic illness between 1st May 2018 and 30th April 2021. *Index of Multiple Deprivation (IMD); Welsh Index of Multiple Derivation (WIMD). The WIMD and the IMD are separate indices with similar indicators but calculated separately for each nation; scores should not be compared across nations.

**Table 3:** Poisson model of the take-up of non-means tested disability benefits (Personal Independence Payment (PIP), Attendance Allowance (AA) and Disability Living Allowance (DLA)) in the last 12 months of life, for all people who died (aged 16+) from chronic illness in England and Wales between 1st May 2018 and 30th April 2021 (for all models n=1,047,911)

|  | Minimally<br>adjusted for age<br>and sex only<br>IRR* [95% CI] | adjusted<br>predicted<br>proportions<br>[95% CI],<br>minimal<br>model¥ | Fully adjusted<br>for all vars<br>IRR* [95% CI] | adjusted<br>predicted<br>proportions<br>[95% CI],<br>full model¥ |
| --- | --- | --- | --- | --- |
| **Sex** |  |  |  |  |
| Female | ref | 66.9 [66.6-67.1] | ref | 64.4 [64.1-64.6] |
| Male | 0.89 [0.89-0.89] | 59.5 [59.3-59.8] | 0.92 [0.92-0.92] | 59.3 [59.1-59.6] |
| **Ethnicity** |  |  |  |  |
| White British | ref | 63.1 [62.9-63.4] | ref | 61.8 [61.6-62.0] |
| Bangladeshi | 1.16 [1.13-1.20] | 73.4 [69.6-77.5] | 1.11 [1.08-1.14] | 68.6 [64.9-72.6] |
| Black African | 0.90 [0.87-0.93] | 56.8 [54.0-59.8] | 0.96 [0.92-0.99] | 59.1 [56.1-62.2] |
| Black Caribbean | 1.03 [1.02-1.05] | 65.2 [63.5-66.9] | 1.05 [1.03-1.06] | 64.6 [63.0-66.3] |
| Chinese | 0.81 [0.77-0.85] | 51.2 [47.8-54.8] | 0.80 [0.76-0.84] | 49.5 [46.1-53.1] |
| Indian | 0.98 [0.96-0.99] | 61.6 [60.1-63.1] | 1.01 [1.00-1.03] | 62.5 [60.9-64.1] |
| Mixed or multiple ethnic groups | 1.04 [1.02-1.07] | 65.8 [63.5-68.3] | 1.06 [1.03-1.08] | 65.2 [62.9-67.6] |
| Other ethnic group | 0.99 [0.97-1.01] | 62.3 [60.3-64.3] | 1.03 [1.01-1.05] | 63.5 [61.4-65.5] |
| Pakistani | 1.11 [1.09-1.13] | 70.0 [67.7-72.4] | 1.06 [1.04-1.08] | 65.3 [63.1-67.7] |
| White other | 0.98 [0.97-0.98] | 61.6 [60.8-62.5] | 0.99 [0.98-1.00] | 61.3 [60.4-62.1] |
| **Area-based deprivation** |  |  |  |  |
| Quintile 1 (most deprived) | 1.19 [1.19-1.20] | 69.7 [69.3-70.1] | 1.13 [1.12-1.13] | 66.1 [65.7-66.5] |
| Quintile 2 | 1.11 [1.10-1.11] | 64.8 [64.4-65.1] | 1.08 [1.07-1.08] | 63.2 [62.8-63.5] |
| Quintile 3 | 1.07 [1.06-1.07] | 62.3 [61.9-62.7] | 1.05 [1.05-1.06] | 61.5 [61.2-61.9] |
| Quintile 4 | 1.04 [1.04-1.04] | 60.7 [60.4-61.1] | 1.03 [1.02-1.03] | 60.2 [59.9-60.6] |
| Quintile 5 (least deprived) | ref | 58.4 [58.0-58.7] | ref | 58.5 [58.2-58.9] |
| **Highest educational qualification** |  |  |  |  |
| No qualifications | ref | 66.5 [66.2-66.8] | ref | 64.6 [64.4-64.9] |
| Apprenticeship | 0.95 [0.94-0.95] | 62.9 [62.2-63.5] | 0.96 [0.95-0.96] | 61.9 [61.3-62.6] |
| Level 1 qualifications | 0.91 [0.90-0.91] | 60.3 [59.7-60.9] | 0.93 [0.92-0.93] | 59.8 [59.3-60.4] |
| Level 2 qualifications | 0.89 [0.88-0.89] | 59.2 [58.6-59.7] | 0.91 [0.91-0.92] | 59.1 [58.6-59.6] |
| Level 3 qualifications | 0.89 [0.88-0.89] | 58.9 [58.2-59.6] | 0.91 [0.90-0.92] | 58.8 [58.1-59.5] |
| Level 4 qualifications and above | 0.85 [0.85-0.85] | 56.5 [56.1-56.9] | 0.88 [0.87-0.88] | 56.7 [56.3-57.1] |
| Other type (Vocational/Foreign) | 0.95 [0.95-0.96] | 63.3 [62.6-63.9] | 0.97 [0.96-0.97] | 62.5 [61.9-63.1] |
| **English/Welsh language proficiency** |  |  |  |  |
| Very well/native speaker | ref | 63.1 [62.8-63.3] | ref | 61.8 [61.6-62.0] |
| Not well/not at all | 1.1 [1.09-1.11] | 69.3 [67.7-71.0] | 1.08 [1.06-1.10] | 66.7 [64.9-68.6] |
| **Region** |  |  |  |  |
| London | ref | 58.7 [58.2-59.2] | ref | 57.7 [57.2-58.2] |
| East Midlands | 1.08 [1.07-1.09] | 63.4 [62.9-63.9] | 1.08 [1.07-1.08] | 62.1 [61.6-62.6] |
| East of England | 1.03 [1.02-1.03] | 60.3 [59.8-60.7] | 1.03 [1.03-1.04] | 59.5 [59.1-60.0] |
| North East | 1.16 [1.15-1.17] | 68.0 [67.3-68.7] | 1.14 [1.13-1.14] | 65.5 [64.8-66.2] |
| North West | 1.14 [1.13-1.15] | 66.9 [66.4-67.3] | 1.12 [1.11-1.13] | 64.6 [64.1-65.0] |
| South East | 1.00 [0.99-1.00] | 58.7 [58.3-59.1] | 1.03 [1.02-1.03] | 59.1 [58.7-59.5] |
| South West | 1.04 [1.03-1.04] | 61.0 [60.5-61.5] | 1.06 [1.05-1.07] | 61.1 [60.6-61.6] |
| Wales | 1.18 [1.17-1.18] | 69.1 [68.5-69.8] | 1.18 [1.17-1.19] | 68.2 [67.5-68.8] |
| West Midlands | 1.09 [1.08-1.10] | 64.0 [63.5-64.5] | 1.08 [1.07-1.08] | 62.1 [61.6-62.6] |
| Yorkshire and The Humber | 1.11 [1.11-1.12] | 65.4 [64.9-65.9] | 1.10 [1.09-1.11] | 63.4 [62.9-63.8] |
| **Underlying cause of death** |  |  |  |  |
| Cancer | ref | 61.9 [61.6-62.2] | ref | 61.8 [61.5-62.0] |
| Dementia | 1.22 [1.21-1.22] | 75.4 [74.9-75.8] | 1.21 [1.20-1.21] | 74.6 [74.2-75.1] |
| Heart failure | 0.84 [0.84-0.85] | 52.1 [51.8-52.4] | 0.83 [0.83-0.84] | 51.5 [51.2-51.8] |
| HIV | 0.73 [0.62-0.85] | 45.0 [36.7-55.2] | 0.74 [0.63-0.86] | 45.8 [37.3-56.1] |
| Liver disease | 0.73 [0.72-0.74] | 45.1 [44.1-46.0] | 0.72 [0.71-0.73] | 44.4 [43.5-45.4] |
| Neurodegenerative diseases | 1.43 [1.42-1.44] | 88.4 [87.4-89.5] | 1.45 [1.44-1.46] | 89.5 [88.5-90.6] |
| Renal failure | 1.05 [1.03-1.07] | 64.9 [63.1-66.8] | 1.03 [1.02-1.05] | 63.7 [61.9-65.6] |
| Respiratory failure | 1.15 [1.15-1.16] | 71.3 [70.6-71.9] | 1.12 [1.11-1.12] | 68.9 [68.4-69.5] |
| Stroke | 0.93 [0.92-0.93] | 57.3 [56.8-57.9] | 0.92 [0.92-0.93] | 56.9 [56.4-57.4] |

\*also adjusted for natural spline of age, see supplementary file, figure 1 for more information
¥predicted proportions are estimated using proportional weights at the mean age of the study population (79 years) and at the average value of categorical variables.

#### Modelling

Benefit take-up increased with age, but the relationship was non-linear and therefore age was modelled as a natural spline with 2 internal knots at percentiles 33 and 66 (corresponding to ages 75 and 86 years, respectively) and boundary knots set to percentiles 10 and 90 (corresponding to age 62 and 93 years, respectively). Supplementary file, figure 2 plots the relationship between the natural spline of age and benefit take-up, adjusted for sex.

Table 3 presents the results for the minimally and fully adjusted models. In the minimally adjusted models, we observed a statistically significant higher take-up rate for women compared to men, for people with no qualifications compared to all other educational groups, and for people with lower English/Welsh language proficiency compared to native or fluent speakers. These associations were attenuated but remained statistically significant in the fully adjusted model.

In the minimally adjusted model, most ethnic minority groups had statistically significantly higher levels of take-up compared to White British people, apart from people with Black African, Chinese, Indian, and White other ethnicity who had significantly lower rates of take-up. In the fully adjusted model, only Black African and Chinese ethnicity was associated with statistically significantly lower take-up rates.

In the minimally adjusted model, higher levels of area-based deprivation were associated with higher levels of take-up in a stepwise gradient, and this relationship remained statistically significant but was attenuated in the fully adjusted model. The predicted proportions from the fully adjusted model show that for people with average age and characteristics, the take-up rate was 66% (95% CI: 66-67%) in the most deprived areas, and 59% in the least deprived areas (95% CI: 58-59%).

In the minimally adjusted and fully adjusted models, the predicted take-up rate for people with cancer remained at 62% (95% CI 62-62%), and this is consistent with the crude rate reported in table 2. Compared to people who died from cancer, those who died from heart failure, HIV, liver disease, and stroke had statistically significantly lower levels of benefit take-up in the minimally and fully adjusted models. People who died from neurodegenerative diseases and those who died from dementia had the highest take-up rates, 90% (95 CI 89-91%) and 75% (74-75%) in the fully adjusted model, respectively. For people who died from dementia, adjusting for age and sex attenuated the take-up rate from the crude rate of 85% (table 2) to 75% (95% CI 75-76%) in the minimally adjusted model.

The analysis exploring the interaction between the main exposures and binary age (working versus pension age) indicates a similar pattern of higher take-up rates among pension age decedents for most subgroups compared to working age, with some notable differences in this pattern according to underlying cause of death (supplementary file, table 4.a-f and figure 4.a-f). For example, among people who died from renal failure, those of pension age had a lower take-up rate, and among people who died from cancer, respiratory failure, dementia, and neurodegenerative disease, there was little difference in the take-up rate between those of working and pension age (supplementary file, table 4.d and figure 4.d).

## Discussion

In this population-based study of all deaths in England and Wales between 1st May 2018 and 30th April 2021, we found that the take-up rate for non-means tested disability benefits in the last year of life among people who died from chronic illness was 66%, indicating a substantial take-up deficit. This means that one in three people who die from chronic illnesses in England and Wales (120,000 people each year) are not receiving the non-means tested benefits to which they are entitled under the ‘Special Rules for End of Life’. After adjusting for sociodemographic factors, we observed wide variation in the take-up rate by underlying cause of death which was lowest for people who died from liver disease (44%), HIV (46%), and heart failure (52%) and highest for people who died from neurodegenerative diseases (90%), and dementia (75%). Across Local Authorities, the age and sex standardised take-up rate varied from 53% to 78% and was generally higher in more deprived areas but with some examples of lower take-up in more deprived areas.

To our knowledge, this is the first study anywhere to report on the benefit take-up rate for people in the last year of life. In the general pension age population, the take-up rate for two low-income (means-tested) benefits (Pension Credit and Housing Benefit for Pensioners) is reported to be 65% and 83%, respectively.^36^ A recent study of claims for working and pension-age disability benefits among people registered blind or partially sighted reported a 76% take-up rate.^18^ In line with this study of blind and partially sighted people, we found that men are less likely than women to receive the benefits to which they were entitled.^18^ However, in contrast to this and other studies^18, 37^, we found that in our study population the take-up rate increased with age. This may partly reflect length of disability and mortality bias, in that older people may have lived for longer with their disability and therefore had more opportunity to claim. We observed the highest take-up rates for people who died from neurodegenerative diseases and dementia. These diseases are associated with a long period of low function and disability which may mean that people have both a greater need, and more time, to claim benefits. Comparatively, deaths from cancer often have a shorter period of low function before death, and deaths from heart failure are characterised by episodic decline in function and an unclear prognosis.^38–40^

The main benefits studied in this analysis, Attendance Allowance (AA) and Personal Independence Payment (PIP), are currently (2026-7) respectively worth up to £5,960 and up to £10,120 (depending on mobility needs) per person, per year. These payments are non-means tested and are intended to help alleviate some of the extra costs of living with serious illness or disability. Receipt of AA or PIP can also enhance entitlement to means-tested benefits for disabled people and carers on lower incomes, acting as a ‘gateway’ benefit to other support. Living with a terminal illness has been estimated to cost households up to £16,000 per year^41^ in lost income and additional out of pocket expenses for travel, medications, equipment, heating and home adaptations.^42–45^ Families living on a low income can spend as much as 98% of their income on these additional costs^46^ with some building up a ‘debt legacy’ to meet the costs.^6^ In our study population overall, 34% of people who died from a chronic illness did not receive the non-means tested benefits to which they were entitled. We do not know how many of these people actively chose not to claim because they did not need the additional financial support, and how many missed out for other reasons. In our analysis, the take-up rate was higher in more deprived areas. However, it is notable that among people who died in neighbourhoods in the most and second-most deprived quintile groups, after adjusting for all other covariates, the predicted non-take-up rate was 34% (95% CI: 34-34%) and 37% (36-37%), respectively (table 3). This is an indication that lack of need for financial support is unlikely to be the only factor driving the take-up deficit.

A lack of awareness of the financial support available, difficulty accessing benefits advice, onerous and intrusive application processes, and stigma surrounding claiming benefits are cited as reasons why people do not take-up the benefits to which they are entitled.^15, 47, 48^ Another barrier specific to claims made under the ‘Special Rules for End of Life’ may be perceived ineligibility due to uncertainty around prognosis, either by the person themselves or by the healthcare professionals responsible for their care. Healthcare professionals are the gatekeepers of access to benefits via the ‘Special Rules’ which provide fast-track access to benefits without the need for further medical assessments. This speeds-up and simplifies the benefit application process significantly, but requires a doctor or specialist nurse to complete, sign and submit an ‘SR1 form’ providing information about their patient’s progressive, incurable disease and stating that the person is likely to have less than 12 months left to live. The SR1 form does not require a specific estimation of prognosis, awards are normally made for 3 years before they are reviewed and the Department for Work and Pensions (DWP) are clear in their guidance that there are no repercussions for healthcare professionals if their patient lives for longer than 12 months.^49^ Yet, qualitative work with healthcare professionals indicates that worry about repercussions is common and prognostic uncertainty remains a key barrier to completing SR1 forms.^50^

Accounts from people living with terminal illness about their experiences of the benefit system highlight other problems including delays to claims, inappropriate withdrawal of benefits, and insensitivity from claim handlers.^41, 51^ Further work to qualitatively understand experiences of, and barriers and facilitators to, making claims under the ‘Special Rules’ is needed to inform improvements to claims processes and take-up initiatives. Our recent review of take-up initiatives for severely disabled people and those with serious, life-limiting illness found a dearth of proactive take-up work among Local Authorities and voluntary and community groups.^52^ Potential models of take-up initiatives include proactive benefit eligibility assessments incorporated into existing processes such as digital advance care planning or General Practitioner palliative care registers, and awareness raising initiatives targeting healthcare professionals working with people with terminal illnesses. Our analysis has identified groups most at risk of under-claiming. Further work is needed to develop and evaluate initiatives to improve take-up overall, with a proportionately greater focus on diagnostic groups and geographical areas with the lowest levels of take-up.

We found that most ethnic minority groups had a higher take-up rate than White British people. Decedents with Bangladeshi and Pakistani ethnicity had the highest rates of take-up, and this was strengthened after adjusting for age and sex and then attenuated after adjusting for other sociodemographic factors and underlying cause of death. After adjusting for all covariates, people of Chinese (49% (95% CI 46-53%)), Black African (59% (56-62%)) and White Other (61% (60-62%)) ethnicity had statistically significantly lower take-up rates compared to White British people (62% (62-62%)). A recent study of benefit take-up found a higher take-up rate among White people when compared to a single aggregate category for non-White ethnic minorities.^18^ Our findings highlight the importance of analysing refined categories of ethnicity where possible, and indicate a need to understand the experiences and preferences of different groups in relation to claiming benefits, in particular people of Chinese ethnicity who have a notably lower take-up than other groups.

## Strengths and limitations

A strength of our study is that we used high quality, highly complete data for the whole population for England and Wales, with 12% of all deaths excluded due to inability to link across datasets. Our analysis focused on people who died from chronic progressive illnesses, where there is a reasonable expectation that a healthcare professional would recognise they were eligible to apply for benefits under the ‘Special Rules for End of Life’. However, it is possible for someone to die suddenly from a chronic illness, for example, after a late diagnosis of cancer, without the opportunity to apply for benefits. A limitation is that we did not have access to other clinical information such as date of diagnosis, comorbidities, or symptom severity that may have explained some of variation in take-up. We used a validated method of grouping leading chronic causes of death^24^, however, underlying cause of death is not always the best way to identify diagnostic groups, for example people with HIV increasingly die from non-AIDS-related conditions^53^ and dementia is often under-reported in mortality data.^54^

A strength of our study is that the large study population and use of refined categories of self-identified ethnicity has highlighted important differences in benefit take-up between ethnic groups not previously reported. Our analysis explored variation in the relationships between exposure variables and benefit take-up for working age compared to pension age decedents. Further intersectional analysis (e.g. investigating how the relationship between diagnosis or language proficiency and take-up rates might vary by ethnic groups) was beyond scope but could be usefully explored in future analysis using similar sources of data with large sample sizes.

Due to the challenges of sharing and linking administrative data between data holding bodies, studies of benefit take-up more commonly combine survey and administrative data to derive estimates of take-up.^16^ This approach is limited by smaller sample sizes but typically generates a richer dataset including information on economic activity and household income.^18^ A limitation of our dataset is that we relied on an area-based measure of deprivation which is subject to the ecological fallacy (i.e. an assumption that everyone in a neighbourhood shares the same socioeconomic profile), and for most of our study population, the area of residence was based on the decedent’s home address at the 2011 Census, with deaths occurring 7 to 10 years later. For people who moved house after the Census, this record will not capture changes in level of area-based deprivation. However, area-based deprivation is included in the individual analysis as a proxy for socioeconomic position, thus its distance from death may be advantageous, since people who move into care homes towards the end of life may have postcodes that poorly reflect their socioeconomic position. In the area-based analysis of the association between take-up and deprivation at LA level, this discrepancy in timepoints for the two measures may be more problematic and should be considered in the interpretation of the results.

Our analysis pre-dates changes to the ‘Special Rules for End of Life’ that have extended eligibility to people who may die within the next 12 months, from the previous 6-month prognosis. In our study population, we observed a gradual increase in the cumulative take-up rate over the last 12 months of life with a steeper increase during the last 6 months of life for working-age people. Future studies could investigate the impact of the changes to the ‘Special Rules’ on the overall level of take-up and for specific groups. Our dataset did not include information on the reason for the claim, so we do not know how many of the claims were made under the ‘Special Rules’. We also do not know whether people were receiving their benefits at the higher rate which is automatically assigned to someone claiming under the ‘Special Rules’. Some people may have been claiming non-means tested disability benefits prior to having a terminal diagnosis. Therefore, while our outcome variable indicates who missed out on claiming benefits under the ‘Special Rules’, it is not a measure of who claimed via that route.

Our analysis demonstrates a method for measuring the take-up of non-means tested benefits in the last year of life that could be used to monitor ongoing trends and evaluate recent changes to the ‘Special Rules’. In the context of a rapidly increasing taxpayer bill for disability benefit payments and constrained public finances, an important area for future study is to understand the impact that access to these benefits has on outcomes and experiences towards the end of life including in bereavement. This could be studied empirically, for example by linking administrative benefit data and health data to investigate the association between benefit take-up and other outcomes relevant to people in the last year of life, such as the number of unplanned hospital admissions and place of death. Qualitative insights are also needed to understand the impact that this financial support has on dignity, independence, and quality of life towards the end of life.

## Conclusion

In England and Wales, approximately 1 in 3 people (120,000 each year) who die from a chronic illness do not receive the non-means tested disability benefits they are entitled to under the ‘Special Rules for End of Life’. The take-up deficit represents a considerable shortfall, potentially undermining the effectiveness of this support. We report wide variation in the level of benefit take-up by underlying cause of death and between areas. Benefit take-up was higher in more deprived areas but substantial proportions of people in the most deprived neighbourhoods did not receive non-means-tested benefits, indicating that lack of need for financial support is unlikely to be the only factor driving the take-up deficit. Our work highlights clinical and socio-demographic groups and geographical areas that could be targeted with proactive take-up initiatives and demonstrates a method for measuring take-up that can be used to monitor future trends.

## Data Availability

All data produced in the present work are available on request to the Office for National Staistics

